# Linking the Heart–Eye–Brain Axis: Ocular and Cerebral Blood Flow After Catheter Ablation in Atrial Fibrillation

**DOI:** 10.1101/2025.08.01.25332610

**Authors:** Nobuhiko Yamamoto, Hideaki Suzuki, Makoto Nakano, Shin-ich Yamanaka, Tomohiro Ito, Hiroyuki Sato, Takahiko Chiba, Kotaro Nochioka, Kentaro Takanami, Masayuki Yasuda, Hiroshi Kunikata, Toru Nakazawa, Satoshi Yasuda

**Affiliations:** Department of Cardiovascular Medicine, Tohoku University Graduate School of Medicine, Sendai, Japan; Department of Diagnostic Radiology, Tohoku University Hospital, Sendai, Japan; Department of Ophthalmology, Tohoku University Hospital, Sendai, Japan

## Abstract

**Aims:** Atrial fibrillation (AF) is associated with cognitive dysfunction even in the absence of clinically apparent brain infarction. This study aimed to investigate whether restoration of sinus rhythm modifies ocular (OBF) and cerebral blood flow (CBF), and to assess the association between changes in these two parameters.

**Methods:** Both of paroxysmal AF (Paf) and persistent AF (PeAF) patients scheduled to recieve catheter ablation (CA) between March 2023 and July 2024 were enrolled. 2days before and approximately 6 months after CA, laser speckle flography (LSFG) to assess the OBF and 99mTc-ethyl cysteinate dimer SPECT to evaluate the CBF were performed. Changes each parameters and the correlation of these changes were analysed among Paf and PeAF, respectively.

**Results:** We enrolled 12 patients each with Paf and PeAF (mean age 67.8 ± 7.2 years; 25% women) without structural heart disease. Mean blur rate (MBR) was used as an OBF index. At baseline, global CBF was lower in PeAF patients in AF rhythm than in Paf patients in sinus rhythm (38.0 ± 2.6 vs 43.4 ± 4.4 mL/100g/min, p = 0.001), while MBR did not differ (9.0 ± 1.5 vs 10.1 ± 2.3, p = 0.186). Following ablation, PeAF patients showed greater increases in MBR and CBF compared to Paf (ΔMBR: +1.5 ± 2.1 vs –0.6 ± 1.8, p = 0.015; ΔCBF: +2.9 ± 2.8 vs –1.1 ± 3.9 mL/100g/min, p = 0.009). A linear mixed-effects model showed a significant positive correlation between changes in MBR and changes in global CBF (p < 0.001).

**Conclusion:** Restoration of sinus rhythm from atrial fibrillation through catheter ablation was associated with improvements in both MBR and CBF, which were positively correlated. These findings suggest a potential interconnection among cardiac rhythm, retinal microcirculation, and cerebral perfusion, supporting the concept of a heart–eye–brain axis.

**What is Known?:** Atrial fibrillation related to cognitive imparements independent on thromboembolistic mechanism.

**What the Sthdy Adds:** Atrial fibrillation reduced both of ocular blood flow and cerebral blood flow, and restoration to sinus rhythm contribute to improving these flows.

Changes in cerebral blood flow were significantly correlated with changes in ocular blood flow.

**Graphical Abstract:** Summary of the study

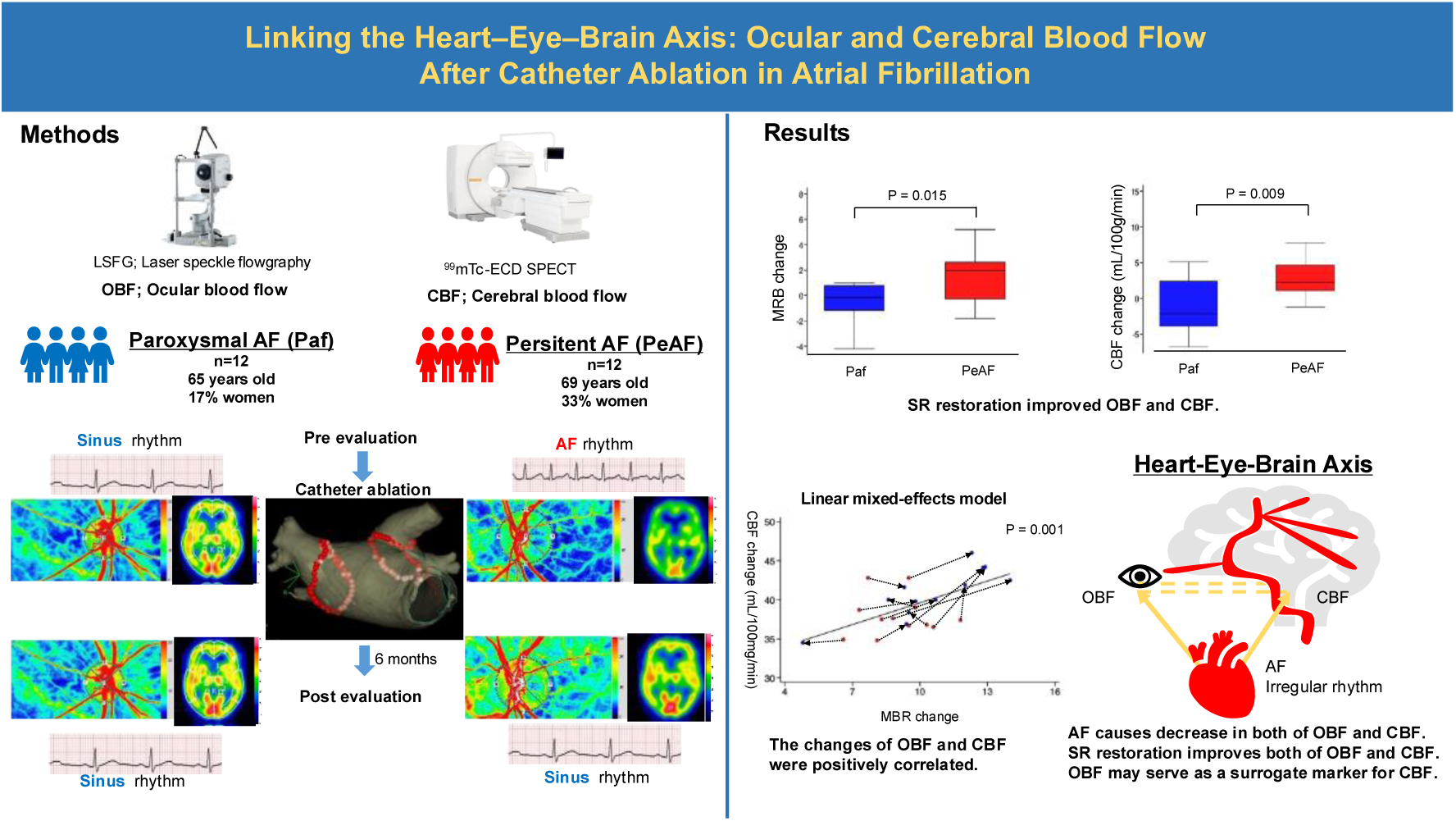

## Introduction

Atrial fibrillation (AF) is increasingly prevalent in aging populations [1–4], and its association with cognitive impairment has become an important clinical concern. The risk of cognitive dysfunction and dementia is elevated in patients with AF even in the absence of clinically evident stroke [5,6]. Among the proposed mechanisms, cerebral hypoperfusion during AF has emerged as a key contributor to cognitive decline, and studies have shown that restoration of sinus rhythm (SR) via catheter ablation (CA) can improve cerebral perfusion [7–14].

Quantitative assessment of cerebral blood flow (CBF) is crucial for understanding the hemodynamic impact of AF. Among available imaging modalities, single-photon emission computed tomography (SPECT) using 99mTc-ethyl cysteinate dimer (ECD) is regarded as a gold standard for noninvasively measuring global CBF due to its high sensitivity and reproducibility [15–19]. However, the clinical utility of SPECT and other imaging techniques such as magnetic resonance imaging (MRI) [10–12,20] is limited by high cost and restricted availability in routine practice [21], highlighting the need for simpler and more accessible alternatives.

Laser speckle flowgraphy (LSFG) has emerged as a promising, noninvasive, and low-cost method to assess ocular blood flow (OBF) [22–26]. This technique evaluates fundus microcirculation by analyzing speckle patterns generated by laser light scattering from moving erythrocytes. The resulting mean blur rate (MBR) serves as an index of retinal blood flow velocity. Since the ophthalmic artery arises directly from the internal carotid artery, which supplies the anterior and middle cerebral arteries, OBF measured by LSFG has been proposed as a potential surrogate for CBF, particularly in perioperative settings [27].

Our group previously demonstrated that LSFG can detect alterations in ocular microcirculation between AF and SR states [28]. However, the direct relationship between OBF and CBF has not been fully elucidated. This study aimed to test the hypothesis that changes in CBF following SR restoration are correlated with simultaneous changes in OBF, by performing direct comparisons using both LSFG and 99mTc-ECD SPECT.

## Methods

### Study population

This prospective observational study consecutively enrolled 14 patients with paroxysmal atrial fibrillation (Paf) and 14 with persistent atrial fibrillation (PeAF), all scheduled to undergo catheter ablation (CA) at Tohoku University Hospital between April 2023 and July 2024. Follow-up was completed by January 2025. Two patients from each group were excluded due to consent withdrawal or loss to follow-up, resulting in a final cohort of 12 Paf and 12 PeAF patients. All participants were free of structural heart disease, as confirmed by cardiac computed tomography, transthoracic echocardiography, and resting electrocardiography. Exclusion criteria included a history of neuropsychological disorders, inability to undergo brain SPECT imaging, failure to provide written informed consent, and ocular diseases precluding retinal evaluation. The study was approved by the Institutional Review Board of Tohoku University (2022-1-353), and written informed consent was obtained from all participants.

### Catheter ablation procedure

All patients received oral anticoagulation with warfarin or a direct oral anticoagulant (DOAC) for at least one month prior to CA, which was continued for a minimum of three months post-procedure. CA was performed under general anesthesia with propofol and dexmedetomidine. Pulmonary vein isolation (PVI) was achieved in all patients using cryoballoon or radiofrequency (RF) ablation under guidance of a three-dimensional mapping system (CARTO3 or EnSite). Additional ablation strategies, including posterior wall isolation, superior vena cava isolation, and ablation of non-pulmonary vein triggers, as well as the choice of energy source, were at the discretion of the operator. Continuation of anticoagulation beyond three months was determined by the treating physician based on thromboembolic risk and AF recurrence.

### LSFG Measurement

Ocular microcirculation was assessed using laser speckle flowgraphy (LSFG; Softcare Co., Ltd., Fukutsu, Japan) within 2 days prior to and at 195.8 ± 26.3 days following CA. The principles of LSFG have been detailed previously [22–24,29]. In brief, LSFG employs a fundus camera equipped with an 830-nm diode laser and a charge-coupled device (CCD) camera to capture the speckle contrast pattern generated by laser light scattered by moving blood cells in the ocular fundus. The mean blur rate (MBR), an index of retinal blood flow velocity, was calculated from this speckle pattern. MBR images were recorded continuously at 30 frames per second over a 10-second period and saved for analysis. Dedicated software synchronized the images with the cardiac cycle and averaged MBR values for each heartbeat, generating a heartbeat-aligned map of the optic nerve head (ONH). The LSFG software automatically segments the optic nerve head (ONH) map into large vessel and capillary areas, providing separate measurements of vessel MBR (mean blur rate in the vessel area), tissue MBR (mean blur rate in the capillary area), and all MBR (mean blur rate in the entire ONH area). In this study, tissue MBR was used for statistical analyses based on the average of three consecutive measurements.

### SPECT image acquisition and CBF measurement

Cerebral blood flow was assessed using 99mTc-ethyl cysteinate dimer (ECD) single-photon emission computed tomography (SPECT), performed 0–1 day before each LSFG measurement. Images were acquired using a dual-head gamma camera system (NM/CT870 DR, GE Healthcare, USA). Global CBF was quantified using the Patlak plot method, as described previously [16].

### Statistical analysis

All statistical analyses were conducted using Stata software version 18 (StataCorp). Continuous variables with normal distribution are presented as means ± standard deviation (SD) and were compared using the Student’s t-test for cross-sectional data. Categorical variables are expressed as percentages and were analyzed using Fisher’s exact test. As no prior studies have reported changes in MBR following CA, sample size estimation was based on differences in global CBF between 6 patients with Paf and 6 with PeAF before CA. A CBF difference of 3.9 mL/min/100 g between the two groups indicated that 12 patients per group would provide 80% power at a two-sided significance level of 0.05. Initial comparisons of CA-induced changes in MBR and global CBF between the Paf and PeAF groups were performed using paired t-tests. Subsequently, multivariable linear regression analyses were used to assess whether changes in MBR and CBF differed significantly between the two groups. Stepwise variable selection was applied with a threshold of P < 0.10 to identify adjustment variables from baseline characteristics that also met this threshold, including heart rate, prior heart failure, estimated glomerular filtration rate (eGFR), left ventricular ejection fraction, left atrial diameter, and brain natriuretic peptide level [30]. Finally, a repeated measures linear mixed-effects model was employed to examine the association between changes in MBR and changes in global CBF [17]. A two-tailed P value <0.05 was considered statistically significant.

## Results

### Baseline characteristics

Baseline characteristics are summarized in **Table 1**. Compared with the Paf group, the PeAF group had significantly higher heart rate (82.4 ± 20.4 vs. 62.8 ± 20.2 bpm, P = 0.027), estimated glomerular filtration rate (eGFR; 75.0 ± 13.7 vs. 58.9 ± 12.3 mL/min/1.73 m², P = 0.017), and brain natriuretic peptide (BNP) levels (147.0 ± 105.4 vs. 28.2 ± 17.4 pg/mL, P < 0.001). No other baseline variables showed significant differences between the two groups (P > 0.05 for all comparisons).

**Table 1.** Baseline Patients Characteristics.

|  | Paf (N=12) | PeAF (N=12) | P value |
| --- | --- | --- | --- |
| Baseline characteristics |  |  |  |
| Age (years) | 65.7±7.5 | 69.8±6.6 | 0.162 |
| Women (%) | 16.7 | 33.3 | 0.640 |
| Body mass index (kg/m<sup>2</sup>) | 25.1±3.1 | 25.1±3.5 | 0.968 |
| Heart rate (bpm) | 62.8±20.2 | 82.4±20.4 | 0.027 |
| CHADS<sub>2</sub> score | 1.4±1.2 | 1.7±1.0 | 0.590 |
| Previous history of heart failure (%) | 8.3 | 50 | 0.069 |
| Hypertension (%) | 66.7 | 66.7 | 1.000 |
| Diabetes (%) | 33.3 | 41.7 | 1.000 |
| Previous history of stroke (%) | 8.3 | 0 | 1.000 |
| eGFR (mL/min/1.73m<sup>2</sup>) | 75.0±13.7 | 58.9±12.3 | 0.017 |
| RAAS inhibitor (%) | 58.3 | 83.3 | 0.371 |
| β blocker (%) | 66.7 | 83.3 | 0.640 |
| MRA (%) | 16.7 | 16.7 | 1.000 |
| SGLT2 inhibitor (%) | 16.7 | 25.0 | 1.000 |
| Left ventricular ejection fraction (%) | 65.2±7.1 | 57.9±11.1 | 0.069 |
| Left ventricular diastolic diameter (mm) | 47.5±3.2 | 47.9±5.0 | 0.811 |
| Left atrial diameter (mm) | 36.8±12.6 | 45.3±4.9 | 0.039 |
| Brain natriuretic peptide (pg/mL) | 28.2±17.4 | 147.0±105.4 | <0.001 |
| Ocular and cerebral hemodynamic measures |  |  |  |
| Mean blur rate | 10.1±2.3 | 9.0±1.5 | 0.186 |
| Global cerebral blood flow (mL/100g/min) | 43.4±4.4 | 38.0±2.6 | <0.001 |

Data are expressed as mean ± standard deviation or percentage. Abbreviations: eGFR,
estimated glomerular filtration ratio; MRA, mineralocorticoid receptor antagonist;
RAAS, renin-angiotensin-aldosterone system; SLGT2, sodium glucose cotransporter 2.

### Changes in OBF and CBF Following CA

Representative LSFG and SPECT images from a patient with PeAF before and after CA are shown in **Figure 1A–B** and **Figure 2A–B**, respectively. Before CA, global CBF was significantly lower in the PeAF group than in the Paf group (38.0 ± 2.6 vs. 43.4 ± 4.4 mL/100 g/min, P = 0.001), while mean MBR did not differ significantly between groups (9.0 ± 1.5 vs. 10.1 ± 2.3, P = 0.186). As shown in **Supplementary Figure 1–2**, CA significantly increased both MBR (from 9.2±1.2 to 10.9±1.7, p=0.029 in the PeAF, and from 9.8±2.5 to 9.1±2.2, p=0.28 in the Paf) and global CBF (from 38.2±2.5 to 41.3±2.7, p=0.005 in the PeAF, and from 43.3±4.4 to 42.1±6.1, p=0.34 in the Paf) in the PeAF group compared to the Paf group. The PeAF group showed greater changes in MBR (1.5 ± 2.1 vs. –0.6 ± 1.8, P = 0.015; **Figure 1C**) and global CBF (2.9 ± 2.8 vs. –1.1 ± 3.9 mL/100 g/min, P = 0.009; **Figure 2C**). These results were consistent in multivariable regression analyses, where PeAF remained significantly associated with changes in both MBR and CBF, with no other variables retained after stepwise selection (**Table 2**). Furthermore, linear mixed-effects modeling demonstrated a significant positive correlation between changes in MBR and global CBF in the PeAF group (P < 0.001; **Figure 3**), whereas no such association was observed in the Paf group (P > 0.05).

**Figure 1:**
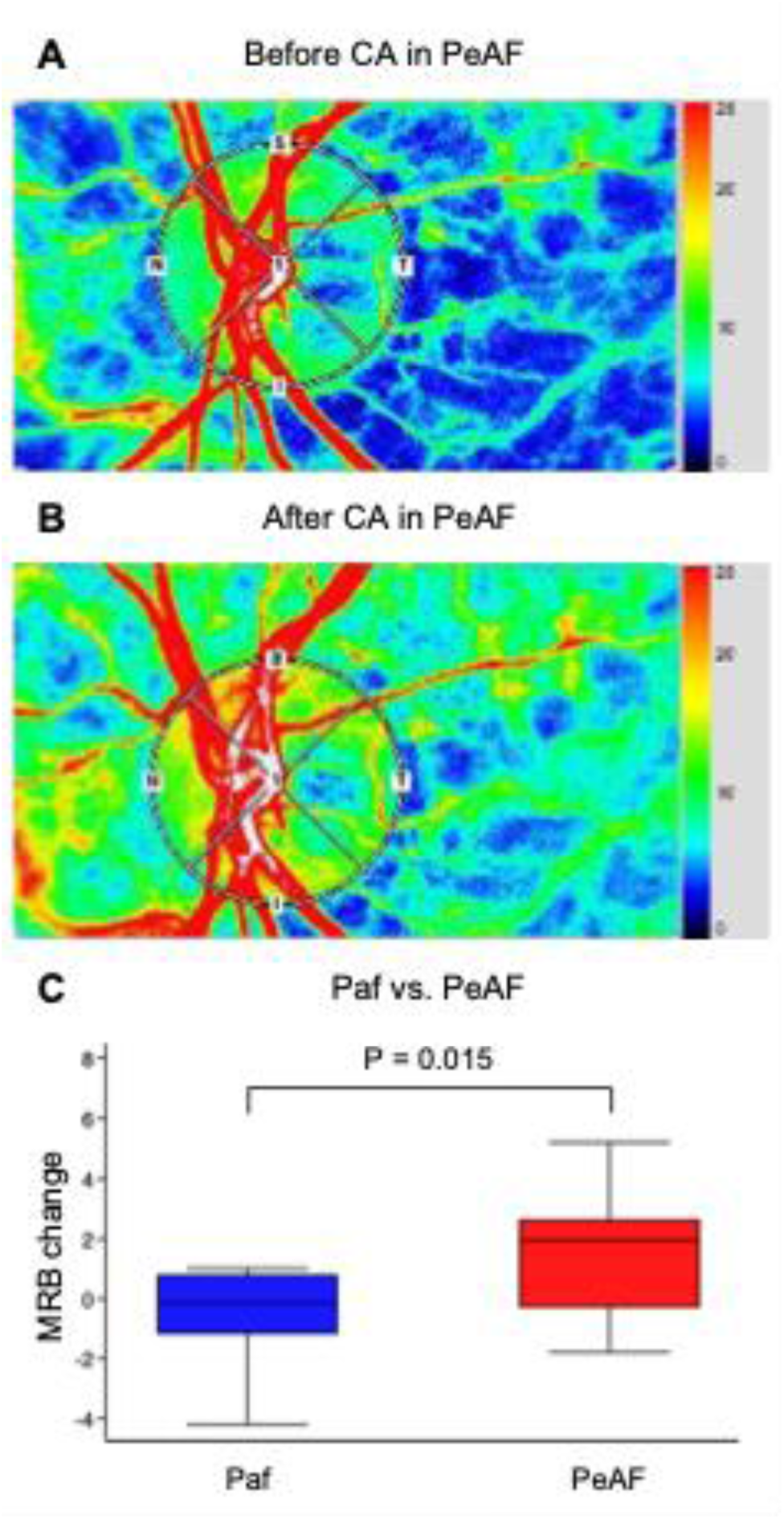
Changes in Ocular Blood Flow Before and After Catheter Ablation. **(A)** Laser speckle flowgraphy (LSFG) image at baseline (before catheter ablation) in a representative patient with persistent atrial fibrillation (PeAF). **(B)** LSFG image at follow-up (after catheter ablation) in the same patient. **(C)** Box plot showing changes in mean blur rate (MBR) before and after ablation in patients with paroxysmal atrial fibrillation (Paf; n=12) and PeAF (n=12). Abbreviations: MBR, mean blur rate; Paf, paroxysmal atrial fibrillation; PeAF, persistent atrial fibrillation.

**Figure 2:**
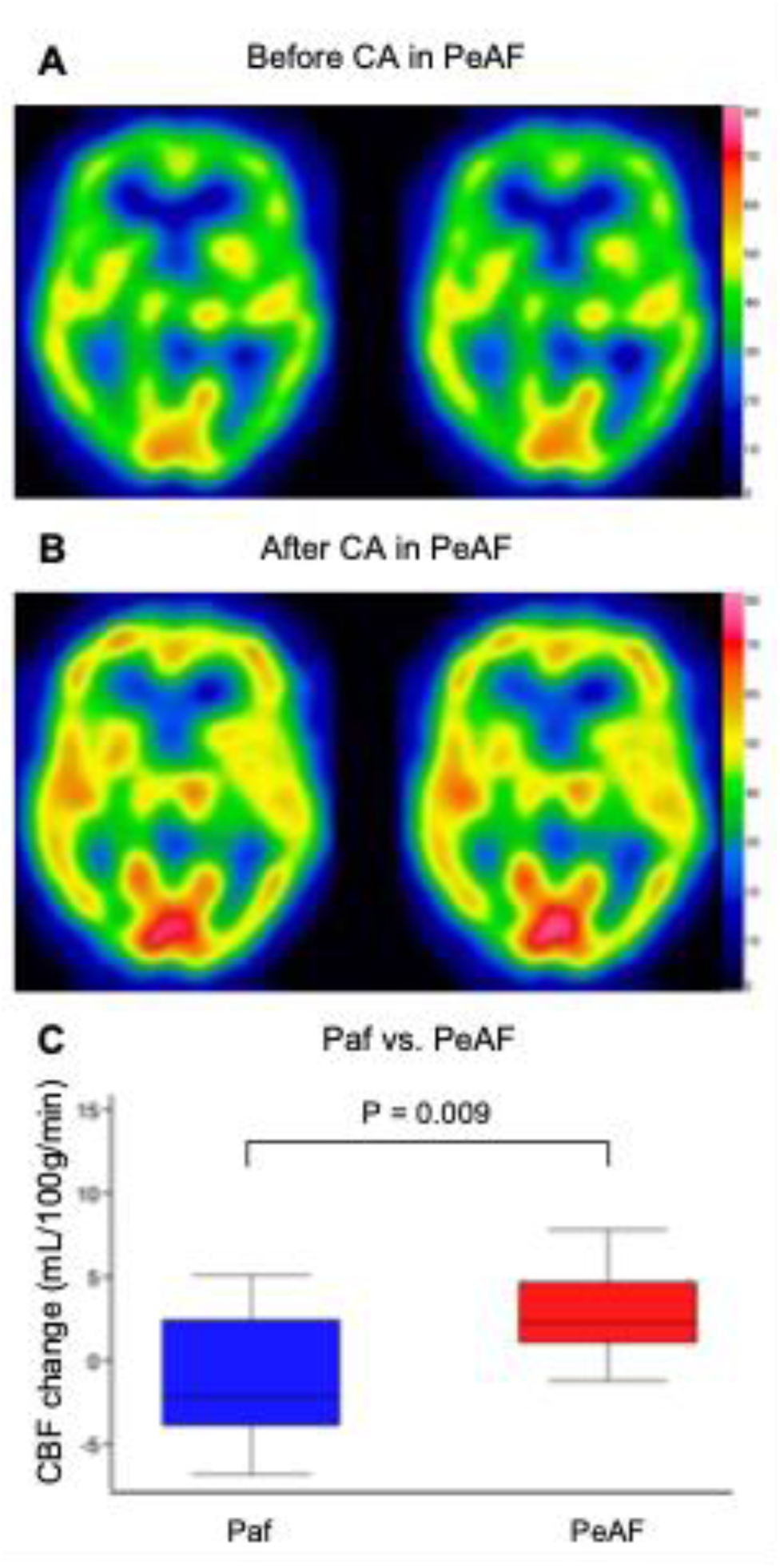
Changes in Global Cerebral Blood Flow Before and After Catheter Ablation. **(A)** Image of cerebral blood flow (CBF) at baseline (before catheter ablation) in a representative patient with persistent atrial fibrillation (PeAF). **(B)** Image of CBF at follow-up (after catheter ablation) in the same patient. **(C)** Box plot showing changes in global CBF before and after ablation in patients with paroxysmal atrial fibrillation (Paf; n=12) and PeAF (n=12). Abbreviations: CBF, cerebral blood flow; Paf, paroxysmal atrial fibrillation; PeAF, persistent atrial fibrillation.

**Table 2.** Multivariable Regression Analysis of Ocular and Cerebral Blood Flow.

|  | Estimate | 95% confidence interval | P value |
| --- | --- | --- | --- |
| MBR |  |  |  |
| Persistent atrial fibrillation | 2.058 | 0.440, 3.676 | 0.015 |
| Global cerebral blood flow |  |  |  |
| Persistent atrial fibrillation | 3.983 | 1.223, 6.844 | 0.009 |

Variables with $P < 0.10$ were selected using stepwise variable selection.
Abbreviations: MBR, mean blur rate

**Figure 3:**
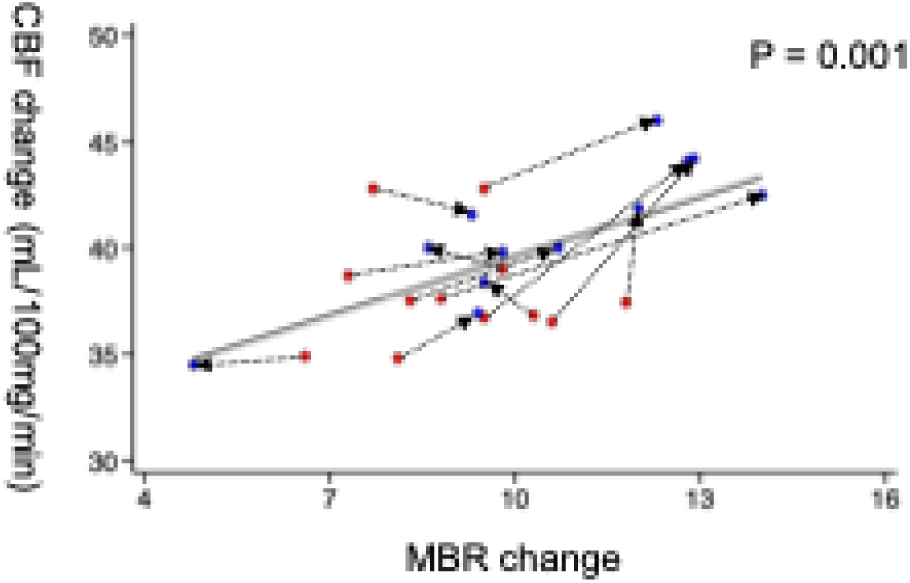
Correlation Between Changes in Ocular and Cerebral Blood Flow in Persistent Atrial Fibrillation. Linear mixed-effects model showing a significant positive correlation between changes in mean blur rate (MBR) and global cerebral blood flow (CBF) in patients with persistent atrial fibrillation (PeAF). Red circles indicate baseline (before catheter ablation) values; blue circles indicate follow-up (after catheter ablation) values. Abbreviations: CBF, cerebral blood flow; MBR, mean blur rate.

## Discussion

This study demonstrated that catheter ablation, a procedure to restore sinus rhythm by electrically isolating arrhythmogenic foci, led to greater increases in both MBR and global CBF in the PeAF group compared with the Paf group. A significant positive correlation between changes in MBR and global CBF was observed in patients with PeAF. This study is the first to directly compare changes in OBF and cerebral blood flow associated with catheter ablation.

### Proposed Mechanism Underlying Decreased CBF and OBF

Dementia is broadly categorized into two major types: vascular dementia, caused by impaired cerebral perfusion due to conditions such as stroke and small vessel disease, and neurodegenerative dementia, such as Alzheimer’s disease, which involves progressive neuronal loss [31]. Vascular factors, however, are known to accelerate the progression of neurodegenerative processes [31–35], and both types often coexist [31].

This study focuses on AF, a common arrhythmia frequently associated with lifestyle-related diseases and hemodynamic impairment. AF has been linked to cognitive decline through multiple pathways, including cerebral infarction [5,8,36], reduced cardiac output [5,37,38], and endothelial dysfunction resulting from inflammatory activation [7,9,13]. These mechanisms are compounded by common comorbidities such as hypertension, diabetes, and chronic kidney disease, all of which contribute to vascular injury and impaired perfusion [33,34,39].

In this study, restoration of SR through CA was associated with increases in MBR and global CBF, indicating enhanced ocular and cerebral perfusion. The high prevalence of hypertension and impaired renal function in the AF population reflects underlying systemic vascular vulnerability, which may contribute to microcirculatory dysfunction [33,39]. Both ocular and cerebral circulations are maintained by autoregulatory mechanisms involving endothelial cells, glial cells, and neurons [25,40]. Chronic vascular risk factors—such as hypertension and kidney dysfunction—can disrupt these mechanisms by impairing endothelial function, diminishing vasodilatory responses, and increasing vascular resistance [41–43]. These findings suggest that restoring SR via CA may improve systemic microcirculation, including OBF and CBF, possibly through modulation of endothelial function. Improved perfusion could in turn help mitigate the risk of cognitive decline in patients with AF.

### LSFG as a Surrogate Marker for CBF Measurement in AF Management

Brain SPECT is an advanced imaging modality that enables the evaluation of cerebral perfusion and metabolism, often detecting abnormalities before structural changes become apparent on brain CT or MRI [44]. Although highly informative, SPECT is costly, time-consuming, and not well suited for repeated use, especially in asymptomatic individuals as part of preventive assessment strategies [45]. In this study, the linear mixed-effects model demonstrated a significant positive correlation between changes in MBR and global CBF, suggesting that OBF assessed by LSFG may serve as a surrogate marker for CBF measured by brain SPECT. Anatomically, the ocular circulation originates from the ophthalmic artery, a direct branch of the internal carotid artery, which also supplies the brain [27]. As such, changes in ocular blood flow may reflect alterations in cerebral circulation. LSFG offers advantages such as simplicity, short examination time, non-invasiveness, and cost-efficiency, making it potentially suitable for repeated assessments and preventive screening. While further validation is required, LSFG may be a practical tool for evaluating cerebral circulation in AF patients undergoing CA.

### Study Limitations

First, subclinical or small brain infarctions may not have been fully excluded, as brain CT or MRI was not performed, and such lesions could be undetectable with 99mTc-ECD SPECT. Second, the influence of heart rate—particularly in PeAF patients prior to CA—may have been underestimated due to the high variability in RR intervals associated with AF. Third, given the relatively short observation period (∼6 months), the long-term effects of sinus rhythm restoration on OBF and CBF remain unclear and warrant investigation through studies with extended follow-up. The linear mixed-effects model accounted for within-subject variability in repeated measures; however, the small sample size may limit the generalizability and stability of the estimates.

## Conclusion

Restoration of sinus rhythm from atrial fibrillation through catheter ablation was associated with improvements in both MBR and CBF, which were positively correlated. These findings suggest a potential interconnection among cardiac rhythm, retinal microcirculation, and cerebral perfusion, supporting the concept of a heart–eye–brain axis in AF management.

## Acknowledgements

The authors wish to thank all ophthalmology and radiology technologists for their technical assistance in the gaining LSFG and brain ^99m^Tc-ECD SPECT data.

## Data availability

The data underlying this article will be shared on reasonable request to the corresponding author.

## Abbreviations

AF: atrial fibrillation
CA: catheter ablation
CBF: cerebral blood flow
LSFG: laser speckle flowgraphy
MBR: mean blur rate,
OBF: ocular blood flow
Paf: paroxysmal atrial fibrillation
PeAF: persistent atrial fibrillation
SPECT: single-photon emission computed tomography,
SR: sinus rhythm

